# Retinal arteriolar tortuosity as a candidate genetic window to inner-ear microvasculature: distributed sharing with blood pressure at CSK and PAWR and a roadmap for larger hearing GWAS

**DOI:** 10.64898/2026.10.08.26365030

**Authors:** Ling Yang, Yang Liu

## Abstract

**Background:** The inner ear is supplied by the labyrinthine artery, which is functionally close to an end artery with limited collateral reserve. Microvascular abnormalities have been implicated in sudden sensorineural hearing loss (SSNHL) and age-related hearing loss, yet inner-ear microvasculature cannot be assessed in vivo and current hearing genome-wide association studies (GWAS) are underpowered. We evaluated the shared genetic architecture of retinal arteriolar tortuosity (retina_AT) with blood pressure, cerebral small vessel disease (cSVD) and hearing phenotypes, and examined whether retinal microvasculature could serve as a candidate genetic window to inner-ear microvasculature.

**Methods:** We used linkage-disequilibrium score regression (LDSC) to estimate SNP heritability for 19 phenotypes and genome-wide genetic correlations (rg) for 11 prespecified pairs. We ran approximate Bayes-factor colocalization (coloc.abf) between 19 independent retinal loci and systolic blood pressure (SBP), together with 95% credible-set fine-mapping, UCSC gene annotation, colocalization with GTEx aortic and tibial-artery expression QTL (eQTL) data and with white-matter hyperintensities (WMH), hearing-GWAS power calculations, and univariable and multivariable Mendelian randomization (MR) of blood pressure and heart rate on hearing.

**Results:** Only retina_AT with SBP was significant genome-wide (rg = 0.10, P = 4.2×10⁻⁵, FDR q = 0.00042). The strongest retinal locus, 13q34, did not colocalize (PP.H3 = 0.996; peaks about 267 kb apart; allelic heterogeneity), whereas two secondary loci colocalized strongly—CSK at 15q24 (PP.H4 = 0.98; 3/3 credible variants within CSK) and PAWR at 12q21 (PP.H4 = 0.99; 3/5 within PAWR)— supported by tissue-matched, moderate eQTL evidence (PP.H4 = 0.71–0.75). At 13q34, retina_AT with WMH was an adequately powered null (PP.H4 = 0.019; only this region and phenotype were tested). On the hearing side, sharing of the blood-pressure magnitude (rg ≈ 0.10) was a near-powered null (about 1.5-fold larger sample needed for 80% power under that assumption, versus about 15-fold for weak rg ≈ 0.03 sharing), and rg ≥ 0.12 was excluded; MR found no robust causal effect.

**Conclusions:** Retinal arteriolar tortuosity shows limited, distributed genetic sharing with blood pressure at the vascular-tone genes CSK and PAWR; no sharing with WMH was observed at 13q34 (only this region was tested). Retinal microvasculature may serve as a candidate genetic window to inner-ear microvasculature, but this analogy awaits direct testing by larger, vessel-oriented hearing GWAS and inner-ear tissue functional studies.

## Introduction

Hearing loss is one of the most common sensory disorders worldwide, and sensorineural hearing loss is its predominant form, among which sudden sensorineural hearing loss (SSNHL) and age-related hearing loss (presbycusis) occupy important positions. The inner ear is supplied mainly by the labyrinthine artery, which is functionally close to an end artery with limited collateral circulation, rendering the cochlea—and especially the stria vascularis—highly sensitive to inadequate microvascular perfusion and dysregulated vascular tone[1]. The resulting “vascular hypothesis” proposes that microvascular spasm, ischemia or endothelial dysfunction can contribute to SSNHL, presbycusis and even noise-induced hearing loss by compromising cochlear oxygen supply and homeostasis. Population-genetic studies have provided some support: SSNHL is associated with inherited and acquired prothrombotic risk factors and various polymorphisms, including MTHFR C677T, prothrombin G20210A, Factor V Leiden, ACE I/D and EDN1[2–5]; animal models likewise link strial microvascular lesions to strial presbycusis[6]. However, these reports derive largely from small case-control, candidate-gene or animal studies with inconsistent results; whether inner-ear microvasculature shares defined biological pathways with systemic vascular abnormalities requires further large-scale population-genetic, cross-disease comparative and functional investigation.

Inner-ear microvascular research is constrained by the deep anatomy and imaging limits of the cochlea: tissue is difficult to obtain, and microvessels and sensory cells measure only 10–50 μm, below the in vivo resolution of MRI, CT and PET, while the blood–labyrinth barrier restricts tracer delivery, leaving the human cochlea a “black box” that cannot be assessed in vivo with high precision[7]. Current hearing GWAS still lack the statistical power to support locus-level colocalization of vascular pathways[8,9]. One of the largest hearing-loss GWAS meta-analyses (17 cohorts; 723,266 participants of European ancestry; 147,997 cases) identified 48 significant loci, with signals enriched in the fusiform, root and basal cells of the cochlear stria vascularis and pointing to a central role for the stria; yet its observed-scale SNP heritability was only h² = 0.0252 (liability scale 0.033–0.061), and much of the genetic architecture remains unexplained by current annotation[8]. A subsequent multi-ancestry GWAS meta-analysis (456,613 cases; 1,053,834 controls) expanded the significant loci to 140 and implicated cochlear hair cells, supporting cells and strial basal and root cells in age-related hearing loss, but SNP heritability remained low[9].

It is therefore of practical value to identify a “surrogate vascular bed” that is directly imageable, genetically tractable and anatomically similar to the inner-ear blood supply[10]. Retinal vessels are an ideal candidate: they are among the few human vascular beds that can be observed directly, non-invasively and at high resolution by fundus imaging[10], and—like cochlear vessels—they share small caliber, limited collateral circulation and high metabolic demand, making both prone to ischemic injury during systemic vascular impairment[10]. Genetically, retinal arteriolar tortuosity is significantly heritable (h² = 0.367, SE = 0.027)[11]; Kirin et al. reported a similarly moderate-to-high heritability in the ORCADES cohort (h² ≈ 0.55, SE = 0.08)[12]. GWAS have identified 89 independent loci (strongest P = 2.00×10⁻¹⁰⁸), supporting a polygenic architecture[11]; the 19q13 locus (ACTN4/CAPN12) is associated with both retinal venular tortuosity and coronary artery disease risk, suggesting shared genetic ground with systemic cardiovascular disease[13]; and the Norrin–Frizzled-4 pathway governs retinal and cochlear vascular development, with defects causing retinal vascular abnormalities and loss of strial vessels[14]. Retinal vascular geometry has also been linked epidemiologically and genetically to blood pressure and cardiovascular risk, and MR indicates that factors related to arterial tortuosity can raise diastolic blood pressure rather than the reverse[11]. Yet a systematic cross-trait dissection of where retinal vascular geometry shares genetic architecture with cSVD or hearing phenotypes is still lacking[10,11].

Accordingly, this study integrates publicly available GWAS summary data across 19 phenotypes—retinal vascular geometry, blood pressure, resting heart rate, cSVD and hearing—using a multilayer framework of genome-wide genetic correlation, locus-level colocalization, fine-mapping and gene annotation, eQTL integration, power quantification and Mendelian randomization, to: (1) characterize the genome-wide and locus-level sharing of retinal arteriolar tortuosity with blood pressure and other traits; (2) identify and annotate candidate causal genes at shared loci; (3) determine whether this sharing extends to cSVD or hearing; (4) quantify the power gap of current hearing GWAS; and, on this basis, evaluate the feasibility of retinal microvasculature as a candidate genetic window to inner-ear microvasculature, providing a roadmap for subsequent vessel-oriented hearing-genetics research.

## Materials and Methods

### Study design

This is a cross-organ genetic-epidemiological study based on publicly available GWAS summary statistics; the overall design and data flow are shown in Fig 1. The work is organized bottom-up into a data-source layer, a primary-analysis layer (LDSC, colocalization, fine-mapping and gene annotation, eQTL colocalization and Mendelian randomization) and a translational-framework layer (hearing-GWAS power analysis and proposal of the candidate retinal window).

**Fig 1.**
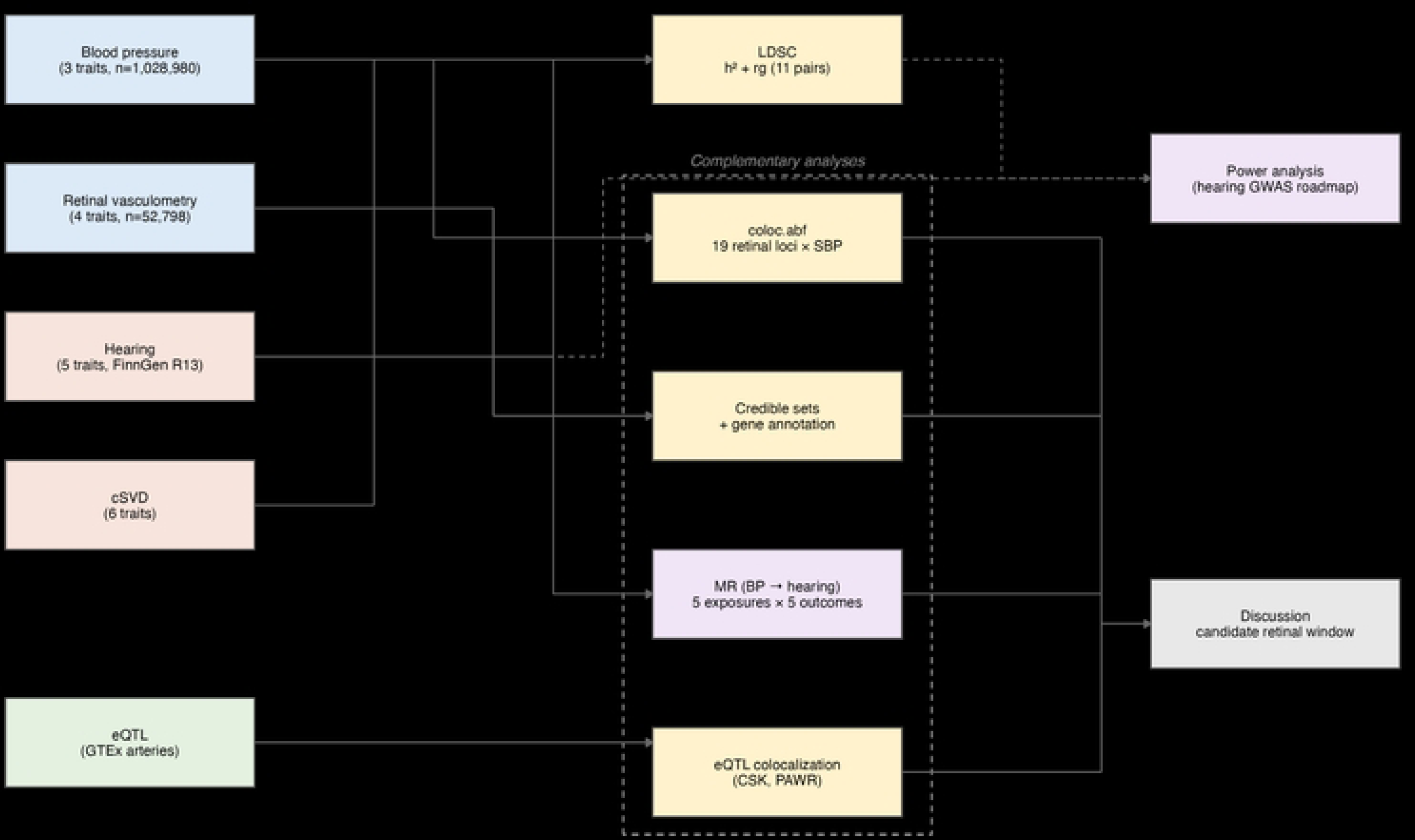
Study design and overall analytical framework. Publicly available GWAS summary statistics for 19 phenotypes (retinal vascular geometry, blood pressure, resting heart rate, cerebral small vessel disease and hearing) enter a bottom-up pipeline comprising a data-source layer, a primary-analysis layer (LDSC heritability and genetic correlations, locus-level colocalization, fine-mapping and gene annotation, eQTL colocalization and Mendelian randomization) and a translational-framework layer (hearing-GWAS power analysis and the candidate retinal-window concept).

### Data sources

We included four retinal vascular geometry traits, three blood-pressure traits, one resting-heart-rate trait, five hearing phenotypes and six cSVD-related traits, and used aortic and tibial-artery eQTL data. Retinal vascular geometry came from Jiang et al. (PLoS Genet, 2023; n = 52,798; GRCh37)[11]; blood pressure from Keaton et al. (Nat Genet, 2024; n ≈ 1,028,980)[15], including SBP, diastolic blood pressure (DBP) and pulse pressure (PP = SBP − DBP); resting heart rate from van de Vegte et al. (Nat Commun, 2023; n = 832,965)[16]; hearing phenotypes from FinnGen release R13[17], including sensorineural hearing loss (SEN_NAS), presbycusis (PRESBY), idiopathic hearing loss (IDIOP), conductive hearing loss (CON_NAS) and the negative-control trait myopia (MYOPIA); and cSVD traits comprising WMH[18], perivascular spaces (PVS)[19], lacunar stroke[20] and cerebral microbleeds (CMB)[21]. eQTL data were obtained for GTEx v8 aorta and tibial-artery tissues through the eQTL Catalogue[22,23]. Sources, accession numbers, sample sizes, ancestry, genome builds and phenotype definitions are detailed in S2 Table.

Regarding sample overlap: all 52,798 participants of European ancestry in the retinal vascular GWAS come from the UK Biobank (UKB); the blood-pressure GWAS (Keaton et al., 2024) is a meta-analysis integrating UKB, ICBP, MVP and BioVU, of which about 24% is from UKB, so there is partial UKB-derived sample overlap between the retinal and blood-pressure GWAS. The hearing GWAS comes from the Finnish FinnGen cohort, an independent Finnish population cohort without UKB participants, and is essentially non-overlapping with the retinal and blood-pressure GWAS. To quantify the impact of overlap, we report the cross-trait LD intercept for retina_AT × SBP as 0.009 (SE 0.0096), not significantly different from zero (z ≈ 0.94, P ≈ 0.35), indicating that, after conditioning on LD score (polygenic signal), there is no detectable residual sharing between the two GWAS attributable to sample overlap or stratification and that rg was not materially inflated by overlap[24]; locus-level colocalization distinguishes shared causal variants through posterior probabilities, and overlap mainly affects the covariance of associations without altering the interpretation of PP.H4[25]. For the two-sample blood-pressure-to-hearing MR, the exposure (UKB/ICBP/MVP/BioVU) and outcome (FinnGen) cohorts do not overlap, satisfying the two-sample assumption[26]. Individual-level de-overlap analysis, as the most stringent validation, is listed as a future direction.

Of note, retinal arteriolar tortuosity used the summary file released by the GWAS Catalog for Jiang et al. (2023) (study accession GCST90270397; n = 52,798; European ancestry); all genetic-correlation, colocalization and heritability estimates in this study are based on this Catalog file, whose locus P values and SNP heritability differ in version/caliber from the values reported in the main text of the original paper, without affecting our conclusions.

### Data preprocessing and quality control

Each GWAS was standardized with munge_sum_stats.py, retaining only variants overlapping HapMap3 with unambiguous allele orientation, and filtering out low-frequency variants, variants with low imputation information and those with anomalous sample sizes. For datasets with nonstandard variant identifiers (such as WMH), chromosome:physical-position was used as the primary matching key; as WMH effect-allele frequencies were entirely missing, the corresponding retinal variant frequencies were used as a proxy for the minor-allele frequency.

### Heritability and genome-wide genetic correlations

LDSC was used to estimate SNP heritability for each phenotype and genome-wide rg between phenotypes. Continuous traits report observed-scale h² directly; for the binary FinnGen hearing phenotypes, in addition to observed-scale h², we transformed to the liability scale using h²_liab = h²_obs · P(1 − P)/z² (P = sample prevalence = cases/total sample; z = standard-normal density at the threshold t = Φ⁻¹(1 − P)). The primary analysis used sample prevalence; SEN_NAS was additionally examined with WHO population prevalence (K = 0.055, 0.076, 0.15) using the standard conversion that distinguishes sample prevalence P from population prevalence K, h²_liab = h²_obs · K²(1−K)²/[P(1−P)·z_K²], which gave 0.062–0.085[27]. Heritability screening was first performed across 19 phenotypes, after which rg was estimated for 11 prespecified pairs with Benjamini–Hochberg FDR correction, defining a positive result as P < 0.05 after correction. Because LDSC rg is the ratio of genetic covariance to genetic standard deviation and the h² scale cancels in this ratio, rg is unaffected by liability-scale transformation.

### Locus-level colocalization

coloc.abf was used to test whether retina_AT and SBP at a given locus are driven by the same causal variant. LD clumping of retina_AT (P < 5×10⁻⁸, r² < 0.05, spacing about 1 Mb) yielded 19 independent lead SNPs. Colocalization windows were centered on the lead SNP: the single-region primary retina_AT × SBP analysis used ±500 kb, and the all-locus analysis across the 19 leads (Table 3) used ±1000 kb to ensure complete coverage of each trait’s signal in the region; for 13q34, where the two peaks are far apart (retinal and SBP peaks 266.7 kb apart), window sensitivity analyses were additionally run at ±250, ±500, ±1000 and ±2000 kb. Priors were p1 = p2 = 1×10⁻⁴ and p12 = 1×10⁻⁵, with PP.H4 > 0.8 defining strong colocalization; p12 was also varied from 1×10⁻⁷ to 1×10⁻⁴ in sensitivity analyses.

### Fine-mapping, credible sets and gene annotation

For strongly colocalized loci, 95% credible sets were constructed from SNP.PP.H4 and assigned to genes through the UCSC Genome Browser; the SuSiE method was used to verify signal independence and to confirm whether the single-causal-variant assumption of coloc.abf held.

### eQTL colocalization

For CSK, PAWR and the neighboring candidate PPP1R12A, we used GTEx v8 cis-eQTL data (via the eQTL Catalogue; GRCh38 coordinates; matched by rsID) from aorta (n = 387) and tibial artery (n = 584), colocalized separately with retina_AT and SBP.

Regarding verification of the single-causal assumption: coloc.abf assumes by default only one causal variant per region. The eQTL Catalogue computes and provides SuSiE credible sets for each dataset; in the CSK and PAWR regions, these SuSiE credible sets did not detect multiple independent eQTL signals, indicating a single association signal in each region and supporting the colocalization obtained under the coloc.abf single-causal assumption. On the GWAS side, the SNP.PP.H4 credible sets at both loci likewise converged into a single signal cluster, with no evidence requiring multi-signal colocalization (coloc.susie).

### Cerebral small vessel disease colocalization

retina_AT and WMH were colocalized in the 13q34 region, with p12 sensitivity analyses to assess robustness.

### Hearing-GWAS power analysis

Based on the variance–sample-size relationship of LDSC genetic-correlation estimates (SE ∝ 1/√N), and using the observed retina_AT × SBP rg = 0.10 as the effect anchor, we calculated the sample size required for detection at various thresholds (principally about 80% power, z ≈ 2.8); this anchor represents the conditional scenario of sharing of the same magnitude as blood pressure (rg ≈ 0.10). We also report the 95% confidence interval from the observed retina_AT × SEN_NAS rg = 0.032 (SE = 0.044) and separately calculate the sample required under a weak rg ≈ 0.032 scenario. Other hearing phenotypes were scaled by the heritability ratio, which was computed on the same scale so that the scale factor cancels; the power results therefore do not depend on the h² scale.

### Mendelian randomization

Two-sample univariable MR used genome-wide significant (P < 5×10⁻⁸), clumped (r² < 0.001) SNPs for each blood-pressure trait and for heart rate as instruments, with random-effects inverse-variance weighted (IVW) analysis as the primary method, reporting ORs per 10 mmHg (per unit for heart rate); the F-statistic assessed instrument strength. Multivariable MR estimated the independent effect of pulse pressure after controlling mean arterial pressure and heart rate and reported conditional F-statistics. We additionally performed Steiger directionality, Cochran’s Q heterogeneity, MR-Egger intercept pleiotropy, leave-one-out and robust adjusted profile score (RAPS) analyses, using CON_NAS and MYOPIA as negative controls.

## Results

### Global genetic architecture

SNP heritability for the 19 phenotypes is shown in Table 1. Retinal arteriolar tortuosity had the highest observed-scale heritability (h² = 0.367, SE = 0.027), followed by WMH (0.344), the other retinal vascular traits (0.091–0.161), blood pressure (0.117–0.137) and heart rate (0.115), all well heritable. Hearing phenotypes had generally low observed-scale heritability: 0.026 for SEN_NAS and 0.010 for MYOPIA, while PRESBY, IDIOP and CON_NAS were only 0.003–0.005; after transformation to the liability scale, SEN_NAS, MYOPIA, PRESBY, IDIOP and CON_NAS were 0.075, 0.125, 0.057, 0.043 and 0.046, respectively. For cSVD_Lacunar and cSVD_CMB_any, the LDSC point estimates were negative and heritability was not estimable (NE; see Table 1 footnote).

**Table 1.**
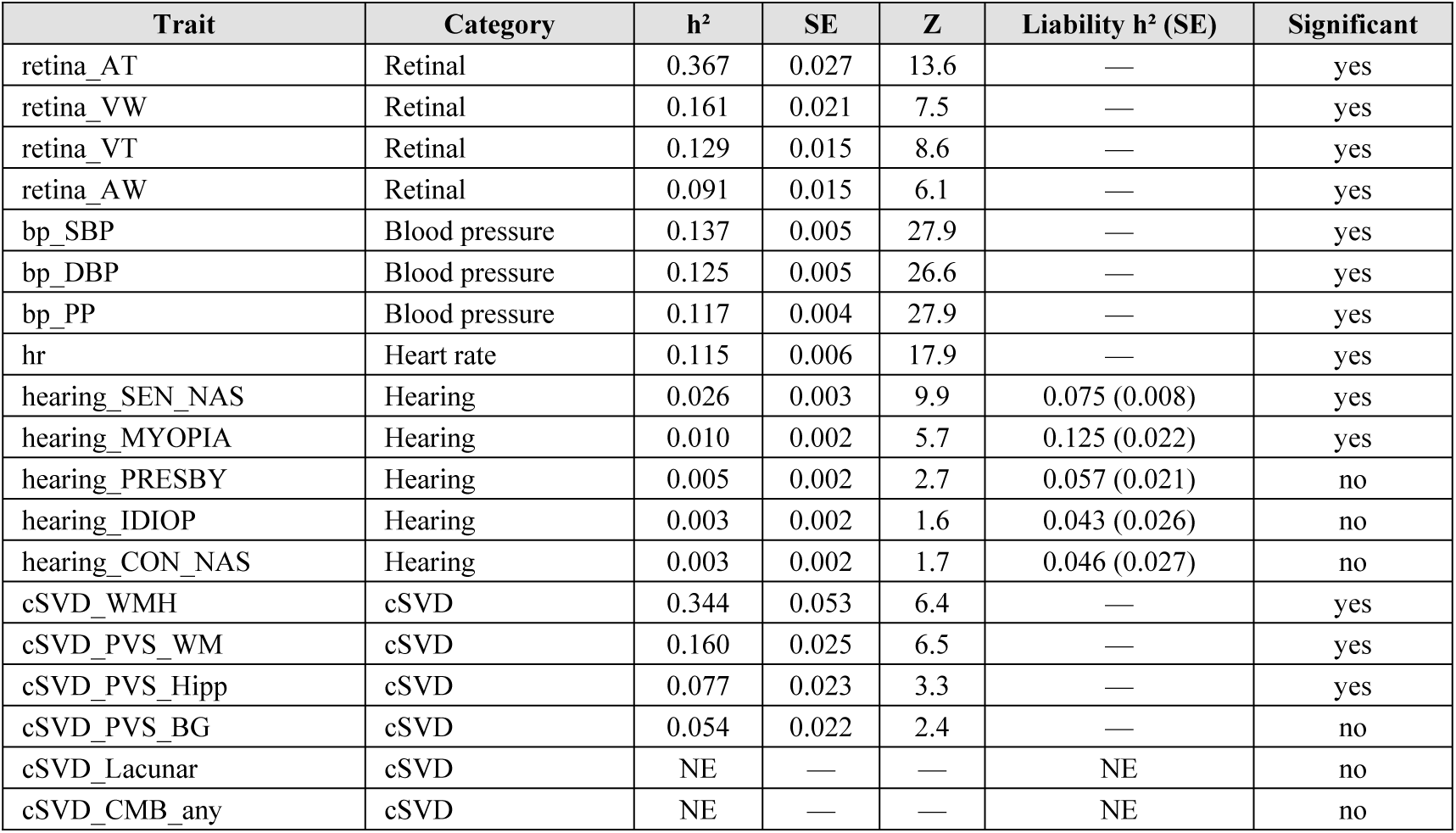
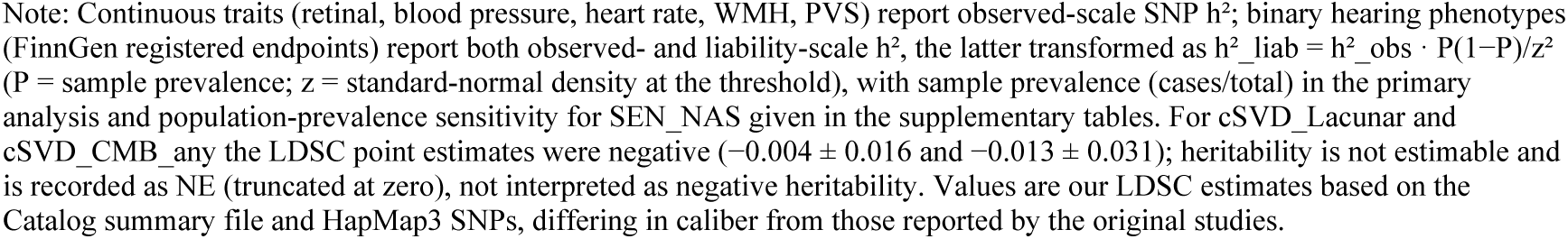
SNP heritability of the 19 phenotypes.

| Trait | Category | $h^2$ | SE | Z | Liability $h^2$ (SE) | Significant |
| --- | --- | --- | --- | --- | --- | --- |
| retina_AT | Retinal | 0.367 | 0.027 | 13.6 | — | yes |
| retina_VW | Retinal | 0.161 | 0.021 | 7.5 | — | yes |
| retina_VT | Retinal | 0.129 | 0.015 | 8.6 | — | yes |
| retina_AW | Retinal | 0.091 | 0.015 | 6.1 | — | yes |
| bp_SBP | Blood pressure | 0.137 | 0.005 | 27.9 | — | yes |
| bp_DBP | Blood pressure | 0.125 | 0.005 | 26.6 | — | yes |
| bp_PP | Blood pressure | 0.117 | 0.004 | 27.9 | — | yes |
| hr | Heart rate | 0.115 | 0.006 | 17.9 | — | yes |
| hearing_SEN_NAS | Hearing | 0.026 | 0.003 | 9.9 | 0.075 (0.008) | yes |
| hearing_MYOPIA | Hearing | 0.010 | 0.002 | 5.7 | 0.125 (0.022) | yes |
| hearing_PRESBY | Hearing | 0.005 | 0.002 | 2.7 | 0.057 (0.021) | no |
| hearing_IDIOP | Hearing | 0.003 | 0.002 | 1.6 | 0.043 (0.026) | no |
| hearing_CON_NAS | Hearing | 0.003 | 0.002 | 1.7 | 0.046 (0.027) | no |
| cSVD_WMH | cSVD | 0.344 | 0.053 | 6.4 | — | yes |
| cSVD_PVS_WM | cSVD | 0.160 | 0.025 | 6.5 | — | yes |
| cSVD_PVS_Hipp | cSVD | 0.077 | 0.023 | 3.3 | — | yes |
| cSVD_PVS_BG | cSVD | 0.054 | 0.022 | 2.4 | — | no |
| cSVD_Lacunar | cSVD | NE | — | — | NE | no |
| cSVD_CMB_any | cSVD | NE | — | — | NE | no |

Note: Continuous traits (retinal, blood pressure, heart rate, WMH, PVS) report observed-scale SNP $h^2$ ; binary hearing phenotypes (FinnGen registered endpoints) report both observed- and liability-scale $h^2$ , the latter transformed as $h^2_{liab} = h^2_{obs} \cdot P(1-P)/z^2$ ( $P$ = sample prevalence; $z$ = standard-normal density at the threshold), with sample prevalence (cases/total) in the primary analysis and population-prevalence sensitivity for SEN\_NAS given in the supplementary tables. For cSVD\_Lacunar and cSVD\_CMB\_any the LDSC point estimates were negative ( $-0.004 \pm 0.016$ and $-0.013 \pm 0.031$ ); heritability is not estimable and is recorded as NE (truncated at zero), not interpreted as negative heritability. Values are our LDSC estimates based on the Catalog summary file and HapMap3 SNPs, differing in caliber from those reported by the original studies.

The genome-wide genetic correlations are shown in Table 2 and Fig 2. Only retina_AT with SBP was significantly positive (rg = 0.10, SE = 0.024, P = 4.2×10⁻⁵, FDR q = 0.00042); retina_AT with heart rate (rg = 0.01, P = 0.68), WMH (−0.07, P = 0.20), white-matter PVS (0.07, P = 0.20) and hippocampal PVS (−0.01, P = 0.93) were not significant, with the comparisons against heart rate and PVS adequately powered and representing powered nulls; retina_AT × SEN_NAS was 0.032 (P = 0.46). retina_VW × WMH (0.116, P = 0.10) showed only a marginal trend. Although retina_AT × SBP was highly significant, rg = 0.10 corresponds to rg² ≈ 0.01, i.e. the two traits share about 1% of genetic variance, indicating that the sharing is real but overall limited. The cross-trait LD intercept for retina_AT × SBP was 0.009 (SE 0.0096), not significantly different from zero, suggesting that sample overlap did not materially contribute to the correlation.

**Table 2.** Genome-wide genetic correlations.

| rg | SE | P | Z | Pair | Retina | Target | FDR | Significant |
| --- | --- | --- | --- | --- | --- | --- | --- | --- |
| 0.10 | 0.024 | 4.2e-05 | 4.1 | retina_AT × bp_SBP | retina_AT | bp SBP | 0.000 | yes |
| 0.116 | 0.071 | 0.10 | 1.6 | retina_VW × cSVD_WMH | retina_VW | cSVD WMH | 0.50 | no |
| 0.073 | 0.057 | 0.20 | 1.3 | retina_AT × cSVD_PVS_WM | retina_AT | cSVD PVS WM | 0.50 | no |
| -0.072 | 0.056 | 0.20 | -1.3 | retina_AT × cSVD_WMH | retina_AT | cSVD WMH | 0.50 | no |
| 0.032 | 0.044 | 0.46 | 0.7 | retina_AT × hearing_SEN_NAS | retina_AT | hearing SEN NAS | 0.84 | no |
| 0.053 | 0.079 | 0.50 | 0.7 | retina_VT × cSVD_WMH | retina_VT | cSVD WMH | 0.84 | no |
| -0.044 | 0.083 | 0.60 | -0.5 | retina_VT × hearing_SEN_NAS | retina_VT | hearing SEN NAS | 0.85 | no |
| 0.010 | 0.025 | 0.68 | 0.4 | retina_AT × hr | retina_AT | hr | 0.85 | no |
| -0.007 | 0.084 | 0.93 | -0.1 | retina_AT × cSVD_PVS_Hipp | retina_AT | cSVD PVS Hipp | 1.00 | no |
| 0.000 | 0.074 | 1.00 | 0.0 | retina_VW × cSVD_PVS_WM | retina_VW | cSVD PVS WM | 1.00 | no |
| NE | — | NE | — | retina_AT × hearing_MYOPIA | retina_AT | hearing MYOPIA | — | no (NE) |

Note: rg, genome-wide genetic correlation; FDR, Benjamini–Hochberg q value. retina\_AT × hearing\_MYOPIA was non-estimable (NE): in the joint rg run its heritability went out of bounds ( $rg = NaN$ ), although the standalone hearing\_MYOPIA $h^2$ estimate was $0.010$ . Of the 11 prespecified pairs, 10 were estimable and are shown with numeric values and one (retina\_AT × hearing\_MYOPIA) is recorded as NE.

**Fig 2.**
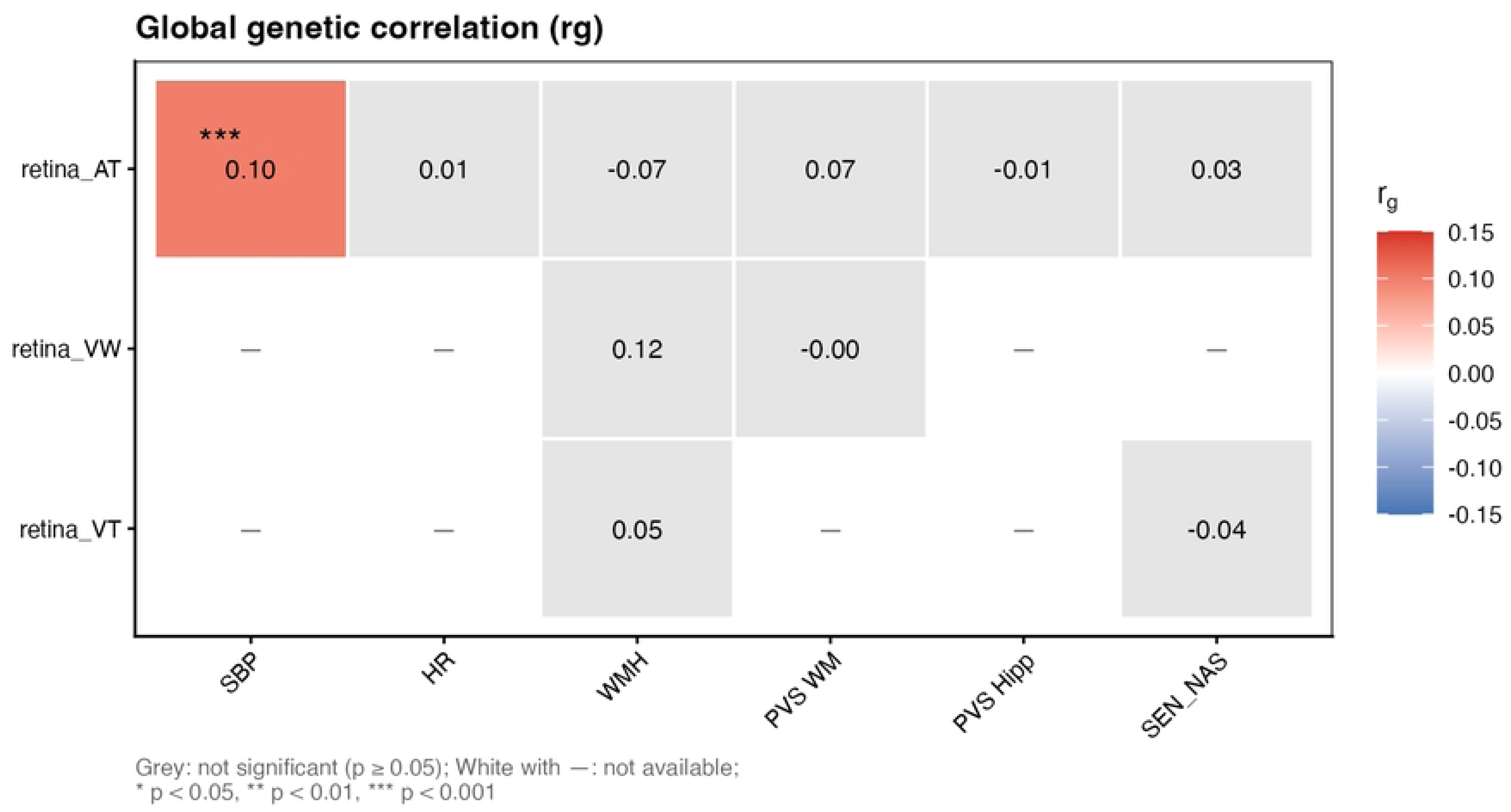
Heatmap of genome-wide genetic correlations. Cells show LDSC rg between retinal vascular traits and blood-pressure, heart-rate, cSVD and hearing phenotypes; only retina_AT × SBP is significant after Benjamini–Hochberg correction (rg = 0.10, q = 0.00042). Color encodes rg direction and magnitude; an asterisk marks the FDR-significant pair. The non-estimable pair retina_AT × hearing_MYOPIA (NE; see Table 2) is not shown; the heatmap displays the estimable comparisons.

### The strongest retinal locus, 13q34, does not share with blood pressure

At the strongest retinal locus, 13q34 (lead SNP rs7991229; retina P = 1.3×10⁻¹⁴²), regional diagnostics showed the retina_AT peak at rs9559797 (P = 7.0×10⁻¹⁵⁴) and the SBP peak at rs9515270 (P = 1.2×10⁻⁸), physically about 266.7 kb apart. Window sensitivity analyses showed that the ±250 kb window, which did not fully cover the independent SBP peak 267 kb away, inflated PP.H4 to 0.183 (PP.H3 = 0.433); once the window fully covered both peaks, ±500 kb gave PP.H3 = 0.964 and PP.H4 = 0.012, ±1000 kb gave PP.H3 = 0.996 and PP.H4 = 0.001, and ±2000 kb gave PP.H3 = 0.994 and PP.H4 = 0.003 (Fig 3). Thus, after window expansion to include the independent SBP peak, the two signals clearly separate, decisively supporting distinct causal variants (allelic heterogeneity; two independent causal variants), and the no-colocalization conclusion strengthens as the window widens.

**Fig 3.**
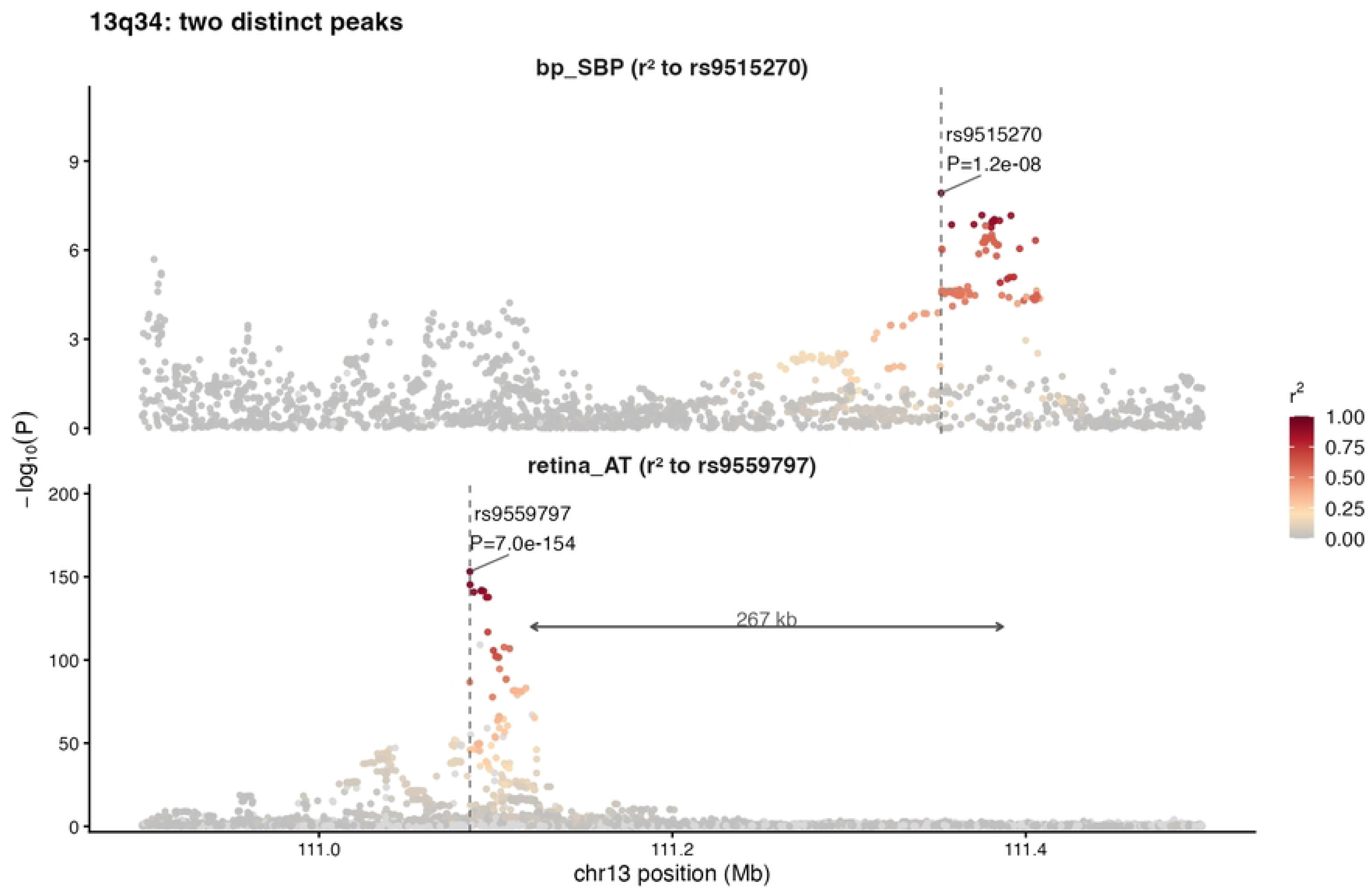
Two independent peaks at the 13q34 locus, about 267 kb apart. Regional association plots show SBP (top) and retina_AT (bottom); the SBP peak (rs9515270; P = 1.2×10⁻⁸) and the retinal peak (rs9559797; P = 7.0×10⁻¹⁵⁴) lie in different LD blocks (r² < 0.01). With windows that fully cover both peaks, PP.H3 ≥ 0.96 and PP.H4 ≤ 0.012, supporting distinct causal variants. Points are colored by LD (r²) with the lead variant.

Complete posterior probabilities across p12 priors for the two shared loci are given in S2 Fig and S3 Table, and those for 13q34 in S3 Table; at 15q24 and 12q21, PP.H4 remained > 0.85 for p12 from 1×10⁻⁶ to 1×10⁻⁴, and the results were robust.

### Two secondary loci, CSK and PAWR, colocalize strongly with blood pressure

Complete colocalization results for the 19 independent retinal loci are shown in Table 3 and S1 Fig. Two secondary loci showed strong colocalization: rs12827748 at 12q21 (PP.H3 = 0.015, PP.H4 = 0.985; posterior odds about 65:1) and rs1378942 at 15q24 (PP.H3 = 0.018, PP.H4 = 0.982; posterior odds about 55:1). rs1035672 showed only weak colocalization (PP.H4 = 0.431), and PP.H4 was very low at all other loci.

**Table 3.** Colocalization of the 19 independent retinal loci with systolic blood pressure.

| Lead SNP | Chr | Position | Retina P | nSNP | PP.H3 | PP.H4 | PO (H4) | Evidence | Anchor gene |
| --- | --- | --- | --- | --- | --- | --- | --- | --- | --- |
| rs12827748 | 12 | 80088578 | 9.7e-28 | 3748 | 0.015 | 0.985 | 65.1 | very strong | PAWR (12q21) |
| rs1378942 | 15 | 75077367 | 1.1e-53 | 3705 | 0.018 | 0.982 | 55.0 | very strong | CSK (15q24) |
| rs1035672 | 2 | 218675199 | 5.3e-73 | 5100 | 0.569 | 0.431 | 0.8 | weak | — |
| rs10415219 | 19 | 39150235 | 4.4e-55 | 5501 | 0.566 | 0.027 | 0 | none | — |
| rs2267406 | 22 | 39633749 | 1.5e-24 | 4353 | 0.954 | 0.013 | 0 | none | — |
| rs10127659 | 1 | 110619665 | 2.1e-32 | 5045 | 0.605 | 0.005 | 0 | none | — |
| rs10163526 | 17 | 68306707 | 5.8e-45 | 5351 | 0.110 | 0.002 | 0 | none | — |
| rs7991229 | 13 | 111091995 | 1.3e-142 | 7420 | 0.996 | 0.001 | 0 | none | Primary locus (excluded) |
| rs12506805 | 4 | 119974540 | 1.1e-45 | 5694 | 0.999 | 0.001 | 0 | none | — |
| rs6833227 | 4 | 8270320 | 4.0e-25 | 7065 | 0.999 | 0 | 0 | none | — |
| rs2147324 | 1 | 150255587 | 3.9e-50 | 2669 | 0.991 | 0 | 0 | none | — |
| rs17086617 | 13 | 28962686 | 1.2e-29 | 6215 | 0.997 | 0 | 0 | none | — |
| rs7683351 | 4 | 85029546 | 5.0e-68 | 4253 | 0.998 | 0 | 0 | none | — |
| rs341917 | 5 | 77949565 | 5.8e-48 | 5177 | 1.000 | 0 | 0 | none | — |
| rs9405366 | 6 | 858296 | 1.1e-21 | 5784 | 1.000 | 0 | 0 | none | — |
| rs3781660 | 11 | 70002689 | 6.5e-31 | 5786 | 1.000 | 0 | 0 | none | — |
| rs2445007 | 8 | 38428657 | 1.3e-24 | 3345 | 1.000 | 0 | 0 | none | — |
| rs9394922 | 6 | 12494502 | 1.3e-24 | 5788 | 1.000 | 0 | 0 | none | — |
| rs10841523 | 12 | 20588575 | 2.7e-59 | 7662 | 1.000 | 0 | 0 | none | — |

Note: PP.H3, posterior probability of distinct causal variants; PP.H4, posterior probability of a shared causal variant; PO (H4), posterior odds for H4. The table uses $\pm 1000$ kb windows centered on each lead SNP (nSNP, number of shared variants in the window); at 13q34 (rs7991229), where the peaks are 266.7 kb apart, additional $\pm 250/\pm 500/\pm 2000$ kb window sensitivity analyses are reported in Section 3.2. PP.H0–PP.H2 were uniformly negligible and are omitted.

The 95% credible sets and gene assignments are shown in Table 4 and Fig 4. At 15q24, the three credible variants (rs1378941, rs2168518, rs1378942) all fall within CSK, establishing CSK as the anchor gene; at 12q21, three of the five credible variants (rs4842267, rs4842318, rs8176822) fall within PAWR and two (rs4842266, rs11614730) lie about 27–53 kb upstream of PAWR, making PAWR the candidate anchor gene. SuSiE verification confirmed a single independent signal at both loci, supporting the coloc.abf single-causal assumption.

**Fig 4.**
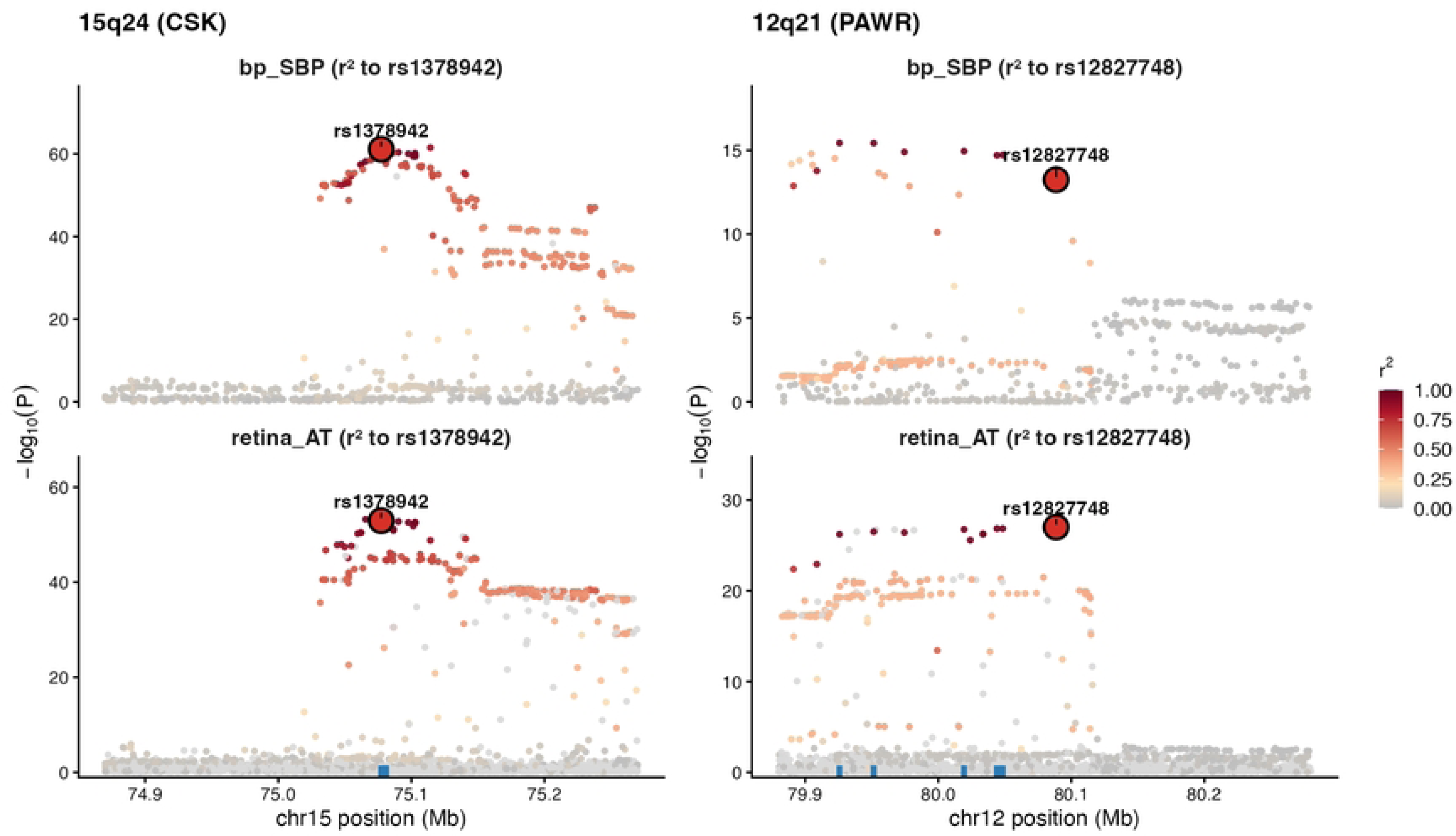
Regional plots of the shared 15q24 (CSK) and 12q21 (PAWR) loci. Association P values (points colored by LD with the lead variant) are shown with annotated genes (GENCODE) below. At 15q24 all three 95% credible-set variants lie within CSK; at 12q21 three of five credible variants lie within PAWR and two lie 27– 53 kb upstream. Both regions colocalized strongly with SBP (PP.H4 = 0.98 and 0.99).

**Table 4.** The 95% credible sets at the two shared loci.

| SNP | Lead SNP | SNP.PP.H4 | Locus | Chr | Position |
| --- | --- | --- | --- | --- | --- |
| rs4842267 | rs12827748 | 0.211 | 12q21 (PAWR) | 12 | 80019427 |
| rs4842266 | rs12827748 | 0.203 | 12q21 (PAWR) | 12 | 79951566 |
| rs11614730 | rs12827748 | 0.185 | 12q21 (PAWR) | 12 | 79925802 |
| rs4842318 | rs12827748 | 0.152 | 12q21 (PAWR) | 12 | 80044167 |
| rs8176822 | rs12827748 | 0.151 | 12q21 (PAWR) | 12 | 80048478 |
| rs1378941 | rs1378942 | 0.389 | 15q24 (CSK) | 15 | 75080150 |
| rs2168518 | rs1378942 | 0.286 | 15q24 (CSK) | 15 | 75081078 |
| rs1378942 | rs1378942 | 0.180 | 15q24 (CSK) | 15 | 75077367 |

Note: SNP.PP.H4, single-variant posterior inclusion probability.

### eQTL colocalization supports a vascular smooth-muscle mechanism

Colocalization of the aortic/tibial-artery eQTLs with the GWAS is shown in Table 5 and Fig 5A. The CSK eQTL in aorta colocalized with retina_AT and SBP (PP.H4 = 0.752 and 0.707), and the PAWR eQTL in tibial artery colocalized with SBP and retina_AT (0.751 and 0.738), all of moderate strength; PAWR in aorta was suggestive (0.54–0.58). CSK in tibial artery (0.069) and PPP1R12A in both tissues (0.036–0.043) did not colocalize, essentially excluding PPP1R12A as a candidate. CSK maps to elastic large arteries and PAWR to muscular arteries, showing a tissue cross-consistency matched to vessel type.

**Table 5.** Colocalization of aortic/tibial-artery eQTLs with the GWAS.

| Tissue | Gene | GWAS | nSNP | PP.H3 | PP.H4 | Tissue match |
| --- | --- | --- | --- | --- | --- | --- |
| Artery–Aorta | CSK | retina_AT | 4720 | 0.140 | 0.752 | Yes |
| Artery–Tibial | PAWR | bp_SBP | 3579 | 0.095 | 0.751 | Yes |
| Artery–Tibial | PAWR | retina_AT | 5101 | 0.120 | 0.738 | Yes |
| Artery–Aorta | CSK | bp_SBP | 3542 | 0.152 | 0.707 | Yes |
| Artery–Aorta | PAWR | retina_AT | 4985 | 0.139 | 0.578 |  |
| Artery–Aorta | PAWR | bp_SBP | 3572 | 0.116 | 0.543 |  |
| Artery–Tibial | CSK | retina_AT | 4819 | 0.310 | 0.069 |  |
| Artery–Tibial | CSK | bp_SBP | 3553 | 0.253 | 0.069 |  |
| Artery–Tibial | PPP1R12A | bp_SBP | 3741 | 0.226 | 0.043 |  |
| Artery–Aorta | PPP1R12A | bp_SBP | 3745 | 0.252 | 0.042 |  |
| Artery–Tibial | PPP1R12A | retina_AT | 5147 | 0.293 | 0.039 |  |
| Artery–Aorta | PPP1R12A | retina_AT | 5008 | 0.313 | 0.036 |  |

Note: PP.H4 of 0.71–0.75 represents moderate supporting evidence.

**Fig 5.**
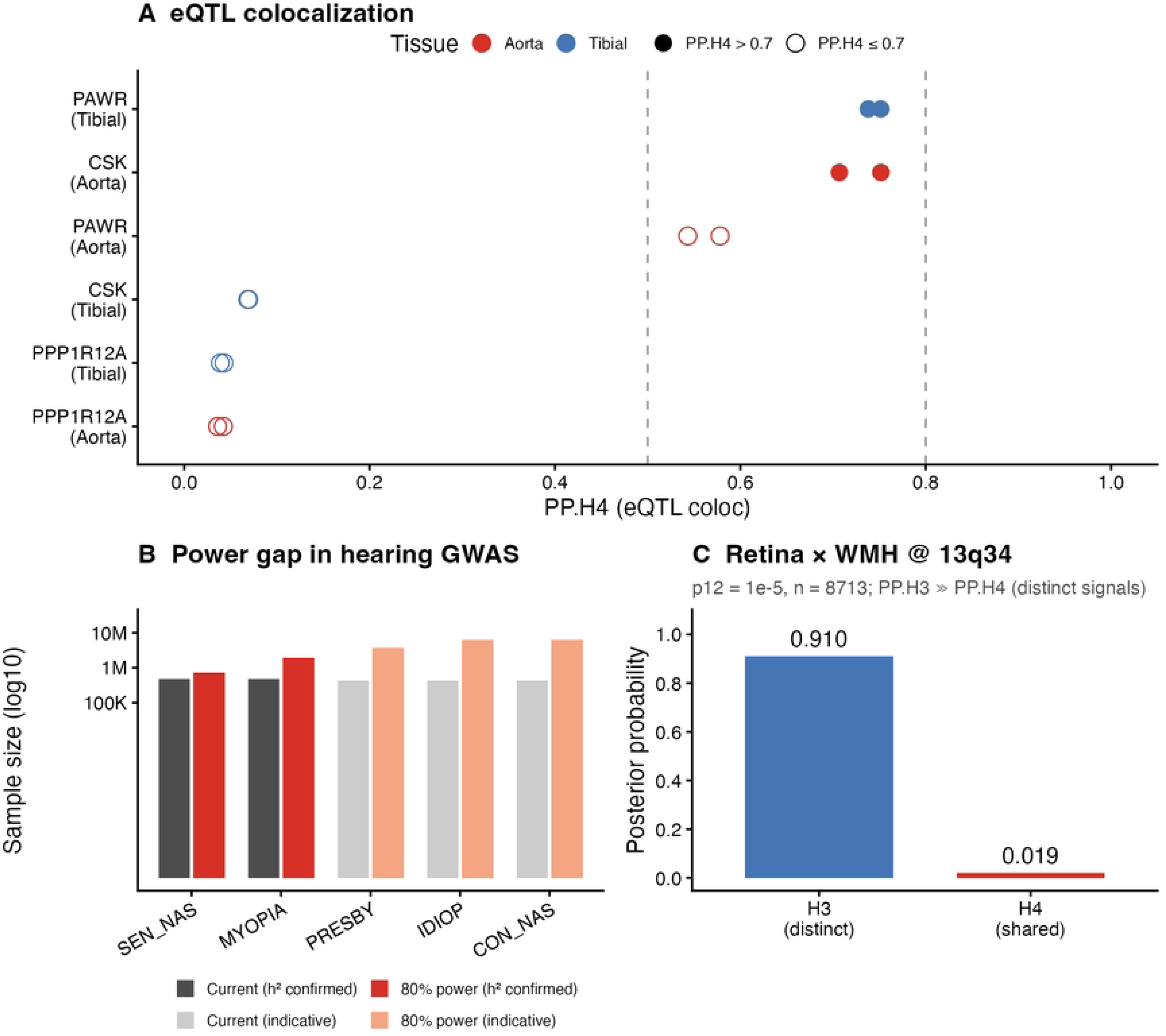
Integrated eQTL colocalization, hearing-GWAS power gap and retina×WMH null. (A) eQTL colocalization: PP.H4 for CSK and PAWR cis-eQTLs (GTEx v8 aorta and tibial artery) against retina_AT and SBP; tissue-matched pairs (CSK–aorta, PAWR–tibial) show moderate support (0.71–0.75), whereas PPP1R12A does not colocalize. (B) Hearing-GWAS power gap: for each hearing phenotype, the current sample size and the sample size required for ≈80% power under an rg ≈ 0.10 (blood-pressure-magnitude) anchor; the weak rg ≈ 0.032 scenario, which requires much larger samples, is described in Section 3.6 and S4 Table. (C) retina_AT × WMH at 13q34: an adequately powered null (8,713 shared variants; PP.H3 = 0.910, PP.H4 = 0.019; only this region and phenotype were tested).

### Sharing does not extend to cerebral small vessel disease

retina_AT and WMH in the 13q34 region included 8,713 shared variants with adequate power, yielding PP.H3 = 0.910 and PP.H4 = 0.019 (Fig 5C), stable in p12 sensitivity analyses (S3 Fig); this is an adequately powered null, indicating that at this locus (13q34) the retina–blood-pressure sharing signal does not overlap WMH. Importantly, we tested only WMH in the 13q34 region, and this cannot exclude sharing at other loci or with other cSVD phenotypes.

### The hearing side is a near-powered null

The observed retina_AT × SEN_NAS rg was 0.032 (SE = 0.044; 95% CI −0.054 to 0.118), excluding stronger sharing of rg ≥ 0.12 but not smaller sharing. The power analysis (Fig 5B; S4 Table) is a conditional inference: if true sharing is of the blood-pressure magnitude (rg ≈ 0.10), about 729,144 participants (about 1.5-fold the current 480,388) are needed for ≈80% power for SEN_NAS; if the true effect is the weak current point estimate rg ≈ 0.032, about 7.12 million (about 14.8-fold) are needed. The hearing-side null should therefore be read as marginal power for blood-pressure-magnitude (rg ≥ 0.10) effects with rg ≥ 0.12 excluded, rather than exclusion of weak rg ≈ 0.03 sharing. Under the rg ≈ 0.10 anchor, MYOPIA, PRESBY, IDIOP and CON_NAS, limited by lower heritability, require about 3.9-, 8.7-, 14.5- and 14.5-fold samples, respectively.

To quantify the current detection capacity of each hearing phenotype, the minimum detectable rg at the present sample size was computed as MDrg = z × SE. At the nominal significance threshold (two-sided z = 1.96), the minimum detectable rg for SEN_NAS, MYOPIA, PRESBY and IDIOP (CON_NAS equal to IDIOP) was about 0.086, 0.139, 0.206 and 0.267; at the 80%-power threshold (z = 2.80), it was about 0.123, 0.199, 0.294 and 0.381 (S4 Table). Using retina_AT × SBP rg = 0.10 as reference, SEN_NAS could detect such an effect at the nominal level but did not reach 80% power, whereas the other phenotypes, with lower heritability and higher minimum detectable rg, had more limited power. This further supports a near-powered null for blood-pressure-magnitude effects on the hearing side and insufficient power for weaker rg ≈ 0.03 effects.

### Blood pressure and heart rate show no robust causal effect on hearing

Univariable MR (random-effects IVW; per 10 mmHg) results are given in S1 Table (panel A): only pulse pressure on SEN_NAS was nominally significant (OR = 0.945, P = 0.038), but it did not pass BH-FDR (q = 0.152); effects of SBP, DBP and MAP on each hearing outcome and of heart rate were not significant (P > 0.18). Multivariable MR (S1 Table, panel B) showed that after adding heart rate, the pulse-pressure coefficient on SEN_NAS attenuated from −0.0074 (P = 0.033) to −0.0033 (P = 0.305), an attenuation of about 56%. Instrument and QC diagnostics are given in S1 Table (panel C): conditional F-statistics were all > 10, Steiger directions were correct in 12 of 12, and the MR-Egger intercept for IDIOP indicated horizontal pleiotropy. Overall, we found no robust causal evidence.

## Discussion

### Principal findings

This study yielded three principal findings through multilayer analysis: genome-wide genetic correlation was observed only between retina_AT and SBP (rg = 0.10, FDR q = 0.00042); this sharing did not occur at the strongest retinal locus, 13q34 (allelic heterogeneity; PP.H3 = 0.996), but was distributed across two vascular-tone genes, CSK at 15q24 and PAWR at 12q21, with tissue-matched, moderate eQTL support; and the sharing did not extend to cSVD in the tested region, the hearing side was a near-powered null, and MR found no robust causal effect.

### Comparison with prior studies

The genome-wide retina–blood-pressure genetic correlation is small in magnitude but highly robust. rg = 0.10 corresponds to rg² ≈ 1%, i.e. the two traits share about 1% of genetic variance, a magnitude comparable to reported genetic-correlation estimates between retinal vascular geometry and other cardiovascular traits. The result is directionally consistent with, and complementary to, the MR finding of Jiang et al. based on the same retinal trait (factors related to tortuosity can influence diastolic blood pressure rather than the reverse): MR indicates the causal direction, whereas the present study delineates the genome-wide extent and specific loci of sharing[11]. Although the genome-wide correlation is highly significant, exclusion of the primary locus 13q34 remains critical, showing that judging cross-organ sharing from the global correlation or the primary locus alone can be misleading.

The 13q34 region contains vascular basement-membrane genes such as COL4A2. Our two strongest tag variants for retina_AT in this region, rs9559797 (P = 7.0×10⁻¹⁵⁴) and rs7991229 (P = 1.3×10⁻¹⁴²), lie only about 6.6 kb apart and are highly linked in 1000-Genomes European populations (r² ≈ 0.90), belonging to one LD haplotype block and representing the same retinal association signal; Ensembl (GRCh37) annotates both within COL4A2. Previous work also identified a retinal arterial tortuosity locus at 13q34 (COL4A2) and verified high COL4A2 expression (and weak COL4A1 expression) in retinal microvascular endothelial cells, suggesting a specific role for COL4A2 in retinal vessels[13]. In contrast, the SBP peak in this region, rs9515270 (P ≈ 1.2×10⁻⁸), lies about 267 kb downstream of the retina_AT peak, is almost unlinked (r² < 0.01), and is annotated outside COL4A2 in the CARS2 gene region; the two peaks belong to different LD blocks and different genes. With colocalization PP.H3 as high as 0.996, this suggests that the primary retinal effect is driven by a COL4A2-related retinal-specific pathway while the blood-pressure signal is driven by an independent neighboring element (CARS2 region), and that cross-trait sharing exists in a distributed form at secondary loci. It should be noted that COL4A1/COL4A2 are also genes implicated in rare-variant cSVD and intracerebral hemorrhage[28]; at the common-variant level we did not observe colocalization of the 13q34 retinal peak with WMH (see Section 3.5). This separation—rare variants causing disease versus common variants regulating—suggests that one cannot directly infer the role of COL4A1/2 common variants in retina–brain sharing from their Mendelian disease associations.

At the cross-trait map level, Veluchamy et al. reported that the 19q13 (ACTN4/CAPN12) retinal venular tortuosity locus is also associated with coronary artery disease risk[13]; the present study instead identifies CSK and PAWR sharing between arterial tortuosity and blood pressure, together delineating a multi-pathway, cross-arterial/venular map of sharing between retinal vascular geometry and systemic cardiovascular disease and indicating that the sharing is locus-specific and distributed rather than dominated by a single major locus. The two shared loci we identify overlap in pathway, but do not coincide fully, with the prothrombotic and endothelial-dysfunction genes emphasized by the previous SSNHL vascular hypothesis. Earlier candidate-gene studies focused largely on the coagulation cascade and endothelial function; for example, hyperhomocysteinemia related to MTHFR C677T/A1298C has been observed in SSNHL, central retinal vein occlusion and cSVD stroke[2], and systematic reviews confirm that endothelial dysfunction—elevated adhesion molecules, imbalanced oxidative stress and reduced brachial flow-mediated dilation— is a potential microcirculatory basis of idiopathic SSNHL[3,5]. CSK and PAWR, however, participate more directly in vascular-tone regulation and smooth-muscle apoptosis: Csk negatively regulates Src-family kinase activity by phosphorylating the C-terminal inhibitory site, and this negative regulation is impaired in vascular smooth-muscle cells from spontaneously hypertensive rats[29], while Par-4 regulates myosin light-chain phosphorylation by binding MYPT1 and participates in vascular smooth-muscle contraction and apoptosis[30]. This suggests that the genetic structure of retina–blood-pressure sharing may operate through a vascular-tone/smooth-muscle-remodeling axis rather than a coagulation/thrombosis axis. This distinction yields a testable prediction: future vessel-oriented SSNHL genetics should be more likely to detect enrichment at tone/smooth-muscle genes than at coagulation genes.

### CSK and PAWR mechanisms

At 15q24, all three credible-set variants (rs1378941, rs2168518, rs1378942) fall within CSK, giving relatively high certainty for the causal candidate. CSK encodes C-terminal Src kinase (Csk), which negatively regulates Src-family kinase activity by phosphorylating the C-terminal inhibitory site and is a key node in vascular endothelial and smooth-muscle signaling, vascular tone and remodeling[31]. In vascular smooth-muscle cells, Csk negative regulation of Src signaling induced by angiotensin II through epidermal growth factor receptor transactivation is impaired in spontaneously hypertensive rats and is considered an upstream mechanism of aberrant ERK1/2 activation[29]. Our CSK aortic eQTL colocalization (PP.H4 = 0.752 and 0.707 with retina_AT and SBP) supports a mechanism in which altered CSK expression changes vascular tone while relating to both retinal tortuosity and blood pressure.

At 12q21, three of five credible-set variants fall within PAWR and two lie about 27–53 kb upstream, making PAWR the candidate anchor gene. PAWR encodes the pro-apoptotic protein Par-4 (PRKC apoptosis WT1 regulator; to be distinguished from the platelet protease-activated receptor PAR4, encoded by F2RL3), participates in vascular smooth-muscle apoptosis and wall-structure regulation, and has been reported as a regulator of myosin light-chain phosphorylation in differentiated contractile vascular smooth muscle[32]. Our PAWR tibial-artery eQTL colocalization (PP.H4 = 0.751 and 0.738 with SBP and retina_AT) is consistent with this. The neighboring candidate PPP1R12A did not colocalize in either tissue (0.036–0.043) and is essentially excluded as a candidate.

The eQTL results also show tissue cross-consistency matched to vascular biology: CSK corresponds to elastic large arteries represented by the aorta, and PAWR to muscular arteries represented by the tibial artery. Elastic large and muscular arteries differ in developmental origin and wall composition (the ratio of elastin to smooth muscle)[33], with the former primarily buffering (the Windkessel effect) and the latter distributing resistance; that the same shared signal anchors to the corresponding gene by vessel type constitutes internal cross-validation difficult to explain by random noise. However, PP.H4 clustered at 0.71–0.75, moderate rather than strong evidence, and GTEx vascular samples are small (aorta n = 387; tibial artery n = 584)[22]; we therefore describe the eQTL results as supportive rather than confirmatory evidence. Colocalization was robust to the p12 prior, with PP.H4 > 0.85 at both loci for p12 from 1×10⁻⁶ to 1×10⁻⁴; sample overlap mainly affects association covariance without altering PP.H4 interpretation[25].

### Interpretation of the null findings

For cSVD, retina_AT and WMH at 13q34 included 8,713 shared variants with adequate power, yielding PP.H3 = 0.910 and PP.H4 = 0.019, stable in p12 sensitivity analyses and representing an adequately powered null. Anatomically, the cerebral circulation has rich anastomoses and strong autoregulation, whereas the retina and inner ear are closer to end-vascular beds; this difference may relate to the null at this locus. Previous work suggests that cSVD may contribute to age-related hearing loss by reducing cochlear blood supply, and WMH has been reported to predict low-frequency hearing loss in older adults independently of age[34]. However, we tested only the single 13q34 region against the single cSVD phenotype WMH, and this cannot imply that sharing genome-wide never involves cSVD; other loci and phenotypes such as PVS, lacunar stroke and CMB remain to be tested.

For hearing, the observed retina_AT × SEN_NAS rg was 0.032 (SE = 0.044; 95% CI −0.054 to 0.118), excluding stronger rg ≥ 0.12 sharing but not smaller sharing. The power analysis is a conditional inference: if true sharing is of the blood-pressure magnitude (rg ≈ 0.10), about 729,144 participants (about 1.5-fold the current 480,388) are needed for ≈80% power for SEN_NAS; if the true effect is the current point estimate rg ≈ 0.032, about 7.12 million (about 14.8-fold) are needed. Under the rg ≈ 0.10 anchor, MYOPIA, PRESBY, IDIOP and CON_NAS require about 3.9-, 8.7-, 14.5- and 14.5-fold samples. The root cause is the generally low SNP heritability of hearing traits: in the largest meta-analysis SEN_NAS observed-scale h² was only 0.0252 (liability scale 0.033–0.061)[8], while our observed-scale estimate of 0.026 is highly consistent and the liability scale was 0.075 with sample prevalence (WHO population-prevalence sensitivity 0.062–0.085), of the same magnitude as the literature liability scale; much of the genetic architecture remains unexplained by current annotation, and although subsequent multi-ancestry GWAS expanded loci to 140 and supported inner-ear (including strial) involvement, SNP heritability remained low[9]. The hearing-side null should therefore be limited to marginal power for blood-pressure-magnitude (rg ≥ 0.10) effects with rg ≥ 0.12 excluded, and does not exclude weak rg ≈ 0.03 sharing; it means weak effects cannot be detected with current samples, not that sharing is absent.

For MR, univariable analysis found only pulse pressure on SEN_NAS nominally significant (OR = 0.945, P = 0.038) but not passing BH-FDR (q = 0.152); multivariable MR showed that after adding heart rate the pulse-pressure coefficient attenuated from −0.0074 (P = 0.033) to −0.0033 (P = 0.305), about 56% attenuation; all other blood-pressure and heart-rate effects were not significant, so no robust causal evidence was found. The exposure (UKB/ICBP/MVP/BioVU) and outcome (FinnGen) cohorts do not overlap, satisfying the two-sample assumption, under which overlap bias is small and tends toward the null[26]. The MR null does not contradict the window hypothesis: the window claims sharing of retinal and inner-ear vascular structure and tone pathways and does not require that blood pressure itself cause deafness. Moreover, the MR null does not exclude a nonlinear, U-shaped relationship between blood pressure and cochlear perfusion (low blood pressure likewise reducing end-bed perfusion, a speculation based on epidemiological observation); MR estimates the lifetime-average effect of instrument variants and cannot capture acute, time-varying perfusion events or the effects of antihypertensive treatment; and FinnGen hearing diagnoses carry coding heterogeneity and misclassification, while the LD structure of the Finnish founder population may affect instrument selection. Observational meta-analyses suggest that hypertension (present versus absent) is associated with about 85% higher risk of hearing loss (OR = 1.85, 95% CI 1.55– 2.21)[35]; these associations may therefore be mediated by nonlinear, local or non-genetic mechanisms or shared confounding rather than a direct causal effect of blood pressure.

### The retina-as-window concept

Based on anatomical parallels, we propose that retinal microvasculature can serve as a candidate genetic window to inner-ear microvasculature, with three layers of evidence. First, anatomical and physiological parallels: the inner-ear labyrinthine artery is functionally close to an end artery with limited collateral reserve, making the cochlea and especially the stria vascularis highly sensitive to tone dysregulation and ischemia yet impossible to observe in vivo[1,7], whereas retinal vessels can be imaged non-invasively, are highly heritable and share small caliber, limited collaterals and high metabolism. Second, developmental homology: Norrin–Frizzled-4 signaling is central to both retinal and cochlear vascular development, and its defects simultaneously cause abnormal retinal vascular development and progressive loss of strial vessels, providing molecular evidence that the two vascular beds share developmental programs[14]. Third, pathway evidence: the CSK and PAWR we identify provide testable candidates that labyrinthine-artery tone may be regulated by the same mechanisms. Clinically, Susac syndrome—characterized by the triad of encephalopathy, branch retinal artery occlusion and sensorineural hearing loss—is a natural example of simultaneous retinal and inner-ear microvascular involvement and can serve as supporting evidence for the window concept; however, its mechanism is immune-mediated microvascular endothelial disease, not equivalent to the tone pathway here[36].

The boundaries and unvalidated nature of this window must be emphasized. This study lacks direct inner-ear molecular evidence; the blood–retinal and blood–labyrinth barriers are not identical in permeability and transport, and the two vascular beds differ in autoregulation and innervation, so this should not be stated as a proven equivalence. To make the hypothesis falsifiable, we explicitly state its test conditions: if larger hearing GWAS still show no sharing at tone loci such as CSK and PAWR, inner-ear (strial/cochlear) tissue eQTL and single-cell data do not enrich for the corresponding pathways, or retinal geometry measures cannot predict hearing outcomes in same-cohort studies, the window hypothesis should be weakened or rejected.

### Translational implications

If the window hypothesis holds, several directions follow. First, fundus imaging is already widely used to screen for diabetic and hypertensive retinopathy, and retinal vascular geometry parameters may become auxiliary predictors of hearing-loss risk: in adults, each 1-SD (18.6 μm) wider retinal venular caliber is associated with 20% higher risk of hearing loss (OR = 1.20, 95% CI 1.03–1.40), and each 1-SD (14.0 μm) narrower arteriolar caliber with 16% higher risk (OR = 1.16, 95% CI 1.00–1.37), with the association significant in adults but not children[37]; each 1-SD increase in retinal arteriolar fractal dimension is associated with a 2.85 dB lower hearing threshold at 1 kHz (β = −2.85, 95% CI −4.47 to −1.23, P < 0.001), and each 1-SD increase in venular fractal dimension with a 2.61 dB lower threshold at 2 kHz (β = −2.61, 95% CI −4.42 to −0.80, P = 0.005), so retinal fractal dimension may serve as a non-invasive biomarker for early detection of hearing loss[38]; tortuosity can reflect the cumulative effect of sustained vascular stress such as hypertension, inflammation and endothelial stress[11]. Second, as tone-regulating genes, the proteins encoded by CSK and PAWR may serve as candidate targets for future vessel-oriented hearing-protective drugs: Csk has been shown to be the causal gene at the 15q24 locus, regulating blood pressure through Src, and both Csk silencing and haploinsufficiency can raise blood pressure[39]; Csk deficiency also causes hypertension by inducing adrenal aldosterone production, increasing sodium reabsorption and plasma volume[40]; but these targets require tissue and functional validation first. Third, our observation that vessel-bed type determines the shared gene (elastic large arteries anchoring CSK; muscular arteries anchoring PAWR) provides tissue-specific clues for subsequent targeting research. These translations require validation in larger, multi-population cohorts and inner-ear functional studies and should not be treated as clinical recommendations at this stage.

### Strengths, limitations and outlook

The strengths of this study lie in its systematic cross-organ phenotype framework, its multilayer evidence chain from genome-wide correlation to gene expression, its power-based distinction of null results, its negative controls and complete QC, and its reproducibility based on public data.

Limitations include: (1) the genome-wide correlation is weak (rg = 0.10; about 1% shared genetic variance) and weak effects may be missed, mitigated by dual LDSC and coloc.abf evidence and FDR control[24]; (2) eQTL support is only moderate and GTEx vascular samples are small, so LD or unmeasured mediation cannot be excluded, described as supportive evidence in light of tissue matching and SuSiE verification[41]; (3) direct inner-ear functional data are lacking, so the analogy cannot be directly tested within this study; (4) the study is predominantly European (Finnish), limiting extrapolation and requiring cross-ancestry validation in East Asian, African and other populations; (5) coloc.abf uses a single-causal assumption and, although SuSiE verification found no multiple signals, may simplify complex regions[25,41]; (6) the retinal GWAS sample size is limited (n = 52,798), affecting the precision of genome-wide correlation, while tortuosity measurement is influenced by image quality, pulsation and other factors, and its reproducibility as a window indicator requires dedicated assessment; and (7) individual identifiers are unavailable in summary data, and there is partial UKB-derived sample overlap between the retinal and blood-pressure GWAS (about 24% of the blood-pressure GWAS from UKB), so individual-level de-overlap sensitivity analysis could not be performed; we quantitatively assessed this with the cross-trait LD intercept (0.009, SE 0.0096, P ≈ 0.35), finding no residual sharing due to overlap; although LDSC, as LD-score regression, is relatively robust to moderate overlap[24] and the two-sample MR has non-overlapping exposure and outcome[26], the direction of residual bias still requires verification in individual data.

Future research should proceed in tiers. Near term: conduct larger hearing GWAS meta-analyses using vessel-oriented phenotype definitions—if the goal is to detect blood-pressure-magnitude (rg ≈ 0.10) sharing, about 729,144 participants are needed for 80% power for SEN_NAS, versus about 7 million for weak rg ≈ 0.03 sharing— and extend colocalization to all retinal loci and multiple cSVD phenotypes such as PVS and lacunar stroke. Medium term: build inner-ear (strial/cochlear) tissue eQTL and single-cell transcriptomes to resolve the cell-type specificity of CSK/PAWR— Trpchevska et al. already mapped GWAS signals to strial fusiform, root and basal cells using mouse cochlear single-cell RNA sequencing[8]—and conduct direct retina–hearing phenotype associations in the same cohorts to test the predictive value of geometry measures. Long term: establish animal models of labyrinthine-artery dysfunction to directly verify the tone mechanism and, on this basis, explore hearing-protection intervention trials targeting the CSK/PAWR pathway.

## Data Availability

All data underlying the findings are publicly available, and no new primary data were generated in this study. The GWAS and eQTL summary statistics used are available from the sources listed below and in S2 Table. Retinal vascular geometry: Jiang et al., GWAS Catalog accession GCST90270397. Blood pressure: Keaton et al. (2024). Resting heart rate: van de Vegte et al. (2023). Hearing phenotypes: FinnGen release R13 (https://r13.finngen.fi/). Cerebral small vessel disease traits: the cited white-matter hyperintensity, perivascular space, lacunar stroke, and cerebral microbleed consortia. eQTL data: GTEx v8 (https://gtexportal.org/) and the eQTL Catalogue (https://www.ebi.ac.uk/eqtl/). Accession numbers, sample sizes, ancestry, genome builds, and phenotype definitions are detailed in S2 Table. All analysis scripts used for data processing, genetic correlation, colocalization, fine-mapping, eQTL colocalization, power analysis, Mendelian randomization, and figure generation are archived at Zenodo via https://doi.org/10.5281/zenodo.23191748. No individual-level data were used, and no restrictions apply to the data analyzed.

## Acknowledgments

We thank the investigators and consortia who made the GWAS and eQTL summary statistics publicly available, including Jiang et al. (retinal vascular geometry), Keaton et al. (blood pressure), van de Vegte et al. (resting heart rate), FinnGen (hearing phenotypes), the cerebral small vessel disease consortia (WMH, perivascular spaces, lacunar stroke and cerebral microbleeds), GTEx and the eQTL Catalogue. The views expressed are those of the authors.

## Author contributions

LY and YL designed the study. LY acquired and curated the data and performed the analyses. LY and YL interpreted the results and wrote the manuscript. YL supervised the work and is the guarantor. Both authors approved the final manuscript and agree to be accountable for all aspects of the work.

## Data availability

All data analyzed in this study are publicly available summary statistics. Sources, accession numbers and download links are listed in S2 Table. The retinal vascular geometry GWAS is from Jiang et al. (GWAS Catalog study GCST90270397); blood pressure from Keaton et al.; resting heart rate from van de Vegte et al.; hearing phenotypes from FinnGen release R13 (https://r13.finngen.fi/); cerebral small vessel disease GWAS from the cited consortia; and eQTL data from GTEx (https://gtexportal.org/) and the eQTL Catalogue (https://www.ebi.ac.uk/eqtl/). All analysis scripts used to process the GWAS and to run the genetic-correlation, colocalization, fine-mapping, eQTL, power and Mendelian-randomization analyses and generate the figures are publicly archived as a Zenodo repository (https://doi.org/10.5281/zenodo.23191748).

## Ethics statement

This study used only publicly available, de-identified GWAS summary statistics and did not involve new human participants or individual-level data; no additional ethical approval was required. Each source study obtained appropriate ethics committee approval and informed consent, as described in the original publications.

## Supporting information

**S1 Fig.** Colocalization of all 19 independent retinal loci with systolic blood pressure.

**S2 Fig.** Posterior probabilities across p12 priors for the two shared loci (CSK and PAWR).

**S3 Fig.** p12-prior sensitivity for retina_AT × WMH at 13q34.

**S1 Table.** Mendelian randomization results: (A) univariable, (B) multivariable, (C) instrument and quality-control diagnostics.

**S2 Table.** GWAS and eQTL data sources, accessions, sample sizes, ancestry, genome builds and phenotype definitions.

**S3 Table.** p12-prior sensitivity posterior probabilities.

**S4 Table.** Hearing-GWAS power analysis and minimum detectable genetic correlations.

**S1 Text.** Supplementary methods and additional analytical details.

